# Comparative effectiveness of sotrovimab versus no treatment in non-hospitalised high-risk patients with COVID-19 in North West London: a retrospective cohort study using the Discover dataset

**DOI:** 10.1101/2023.07.26.23293188

**Authors:** Myriam Drysdale, Evgeniy R. Galimov, Marcus J. Yarwood, Vishal Patel, Bethany Levick, Daniel C. Gibbons, Jonathan D. Watkins, Sophie Young, Benjamin F. Pierce, Emily J. Lloyd, William Kerr, Helen J. Birch, Tahereh Kamalati, Stephen J. Brett

## Abstract

**Introduction:** There is uncertainty regarding how *in vitro* antibody neutralisation activity translates to the clinical efficacy of sotrovimab against severe acute respiratory syndrome coronavirus 2, although real-world evidence has demonstrated continued effectiveness during both BA.2 and BA.5 predominance. We previously reported descriptive results from the Discover dataset for patients treated with sotrovimab, nirmatrelvir/ritonavir or molnupiravir, or patients at highest risk per National Health Service (NHS) criteria but who were untreated. This study sought to assess the effectiveness of sotrovimab compared with no early coronavirus disease 2019 (COVID-19) treatment in highest-risk patients with COVID-19.

**Methods:** Retrospective cohort study using the Discover dataset in North West London. Patients had to be non-hospitalised at index, aged ≥12 years old and meet ≥1 of the NHS highest-risk criteria for receiving early COVID-19 treatment with sotrovimab. The primary objective was to assess the risk of COVID-19-related hospitalisation and/or COVID-19-related death within 28 days of the observed/imputed treatment date between patients treated with sotrovimab and highest-risk patients who received no early COVID-19 treatment. We also performed subgroup analyses for patients aged <65 and ≥65 years, patients with renal dysfunction, and by Omicron subvariant prevalence period (BA.1/2 emergence: 1 December 2021–12 February 2022 [period 1]; BA.2 reaching and at its peak: 13 February–31 May 2022 [period 2]; BA.2 falling and BA.4/5 emergence: 1 June–31 July 2022 [period 3]). Inverse probability of treatment weighting based on propensity scores was used to adjust for measured known and likely confounders between the cohorts. Cox proportional hazards models with stabilised weights were performed to assess hazard ratios (HRs).

**Results:** A total of 599 highest-risk patients treated with sotrovimab and 5,191 untreated highest-risk patients were included. Compared with untreated patients, sotrovimab treatment reduced the risk of COVID-19 hospitalisation or death by 50% (HR=0.50; 95% confidence interval [CI] 0.24, 1.06); however, statistical significance was not reached (p=0.07). In addition, sotrovimab reduced the risk of COVID-19 hospitalisation by 57% (HR=0.43; 95% CI 0.18, 1.00) compared with the untreated group, although also not statistically significant (p=0.051). Among patients aged ≥65 years and patients with renal disease, sotrovimab treatment was associated with a significantly reduced risk of COVID-19 hospitalisation, by 89% (HR=0.11; 95% CI 0.02, 0.82; p=0.03) and 82% (HR=0.18; 95% CI 0.05, 0.62; p=0.007), respectively. In period 1, sotrovimab treatment was associated with a 75% lower risk of COVID-19 hospitalisation or death compared with the untreated group (HR=0.25; 95% CI 0.07, 0.89; p=0.032). In periods 2 and 3, HRs of COVID-19 hospitalisation or death were 0.53 (95% CI 0.14, 2.00; p=0.35) and 0.78 (95% CI 0.23, 2.69; p=0.69), respectively, for the sotrovimab versus untreated groups, but differences were not statistically significant.

**Conclusions:** Sotrovimab treatment was associated with a significant reduction in risk of COVID-19 hospitalisation in patients aged ≥65 years and those with renal disease compared with the untreated cohort. For the overall cohort, the risk of hospitalisation following sotrovimab treatment was also lower compared with the untreated group; however, this did not achieve statistical significance (p=0.051). The risk of hospitalisation and/or death was lower for the sotrovimab-treated cohort across all time periods but did not reach significance for periods 2 and 3.

## Introduction

In March 2020, the rapid global spread of severe acute respiratory syndrome coronavirus 2 (SARS-CoV-2) resulted in the declaration of the coronavirus disease 2019 (COVID-19) pandemic by the World Health Organization.^1^ Some patients are at particularly high risk of severe outcomes from COVID-19, such as those with cancer, renal and liver disease, human immunodeficiency virus/acquired immune deficiency syndrome (HIV/AIDS) and rare neurological conditions.^2, 3^ In England, early treatment of COVID-19 with either antivirals or monoclonal antibodies (mAbs) is recommended for people who meet these ‘highest-risk’ criteria, following approval of these drugs by the United Kingdom (UK) Medicines and Healthcare products Regulatory Agency in late 2021.^4–6^

Sotrovimab is a dual-action engineered human IgG1κ mAb derived from the parental mAb S309, a potent neutralising mAb directed against a conserved epitope in the spike protein of SARS-CoV-2.^7–10^ Intravenous sotrovimab 500 mg was shown in COMET-ICE, a randomised clinical trial, to significantly reduce the risk of all-cause >24-hour hospitalisation or death by 79% compared with placebo in high-risk patients with mild-to-moderate COVID-19.^11^ Sotrovimab received conditional marketing authorisation in December 2021 in the UK for use in symptomatic patients with acute COVID-19 (≥12 years of age and ≥40 kg) who do not require supplemental oxygen but are deemed to be at increased risk of progression to severe COVID-19.^4^ During the study period, the National Health Service (NHS) England clinical guidelines recommended sotrovimab as a first-line treatment option.^3^ The guidelines have since changed to recommend sotrovimab as a second-line option.^12^

Since the COMET-ICE trial was undertaken from August 2020 to March 2021,^11^ new COVID-19 variants of concern have emerged, including the Omicron BA.1 and BA.2 subvariants which became dominant globally in January and March 2022, respectively.^13–15^ *In vitro* neutralisation assays have demonstrated that sotrovimab retained its neutralisation capacity against Omicron BA.1 (3.8-fold IC_50_ reduction relative to wild-type SARS-CoV-2), but showed a moderate reduction against Omicron BA.2 (15.7-fold reduction).^10^ A similar reduction in activity has been reported for BA.5 (21.6-fold reduction), which was predominant in the UK from July to October 2022.^16, 17^

In the absence of clinical trial data, uncertainty remains regarding how *in vitro* antibody neutralisation activity translates to clinical effectiveness, especially for dual-action antibodies like sotrovimab. Despite some emerging real-world evidence demonstrating the effectiveness of sotrovimab versus antivirals during both BA.2 and BA.5 predominance, further evidence is critical for providing up-to-date clinical recommendations when considering the evolving variant landscape.^18^ This study uses the real-world dataset, Discover, to further explore the clinical effectiveness of sotrovimab relative to no treatment for patients in North West London during this time frame. Our previous paper reports descriptive results for patients treated with sotrovimab, nirmatrelvir/ritonavir or molnupiravir, or patients at highest risk per NHS criteria but who were untreated.^19^ Here, we assessed the clinical effectiveness of sotrovimab compared with no early COVID-19 treatment in highest-risk patients with COVID-19 in North West London who did not require initial inpatient management from December 2021 to July 2022.

## Methods

### Study objectives

The primary objective was to assess the risk of COVID-19-related hospitalisation and/or COVID-19-related death within 28 days of the index date (actual or imputed treatment start date) between patients treated with sotrovimab and highest-risk patients who received no early treatment for COVID-19 (untreated patients).

The secondary objectives were to assess the risk of COVID-19-related hospitalisation and/or COVID-19-related death within 28 days between sotrovimab-treated and untreated patients among the following subgroups: Omicron subvariant prevalence period (Additional File 1, Figure S1); patients aged <65 years and ≥65 years at index; and patients with renal dysfunction (‘renal disease’: renal transplant recipients, non-transplant recipients receiving a comparable level of immunosuppression to renal transplant recipients; chronic kidney disease stage 4 or 5).

### Data source and study design

This retrospective cohort study was based on data from the Discover dataset, one of Europe’s largest linked longitudinal datasets.^20^ Discover holds depersonalised coded primary and secondary care data for over 2.7 million patients who are registered with a general practitioner (GP) in North West London. The dataset is fed by data from over 400 provider organisations, including over 350 general practices, two mental health and two community trusts, and all acute providers attended by patients from North West London.^21^ The Discover dataset population has a comparable age–sex distribution and prevalence of comorbidities to the overall UK population, but is more ethnically diverse.^20^ The dataset is accessible via Discover-NOW Health Data Research Hub for Real World Evidence through their data science specialists and Information Governance committee-approved analysts, hosted by Imperial College Health Partners.

In the sotrovimab-treated cohort, the index date was defined as the date of sotrovimab prescription. Patients in the treated cohort must have had a recorded prescription for sotrovimab within 28 days of their COVID-19 diagnosis. In the untreated comparator cohort, index dates were imputed based on the distribution of time to treatment (time from COVID-19 diagnosis date to date of sotrovimab prescription) in the treated cohort (Additional File 1, Figure S2). The baseline period was defined as the 365 days immediately prior to index. Patients were followed up for 28 days from the index date (acute period), during which time patient outcomes were evaluated.

As there were no sequencing data available for patients included in the study, dominance period for Omicron subvariant was used as a surrogate.^22^ Patients were classified into three different variant prevalence periods based on the period in which their diagnosis fell: Omicron BA.1/2 emergence: 1 December 2021 to 12 February 2022 (period 1); BA.2 increasing and at its peak: 13 February 2022 to 31 May 2022 (period 2); BA.2 falling and BA.4/5 emergence: 1 June 2022 to 31 July 2022 (period 3) (Additional File 1, Figure S1).

### Study population

Patients in both cohorts were eligible for inclusion if they were aged ≥12 years on the index date and met at least one of the NHS highest-risk criteria for receiving early treatment with sotrovimab. At the time of study, these criteria included Down’s syndrome, solid cancer, haematological diseases (including cancers), renal disease, liver disease, immune-mediated inflammatory disorders, immune deficiencies, HIV/AIDS, solid-organ and stem-cell transplant recipients and rare neurological conditions.^2, 3^ Patients meeting the NHS highest-risk criteria were identified via the presence of International Classification of Disease version 10 and Systematized Nomenclature of Medicine (SNOMED) codes appearing at any time in the patient’s records since first registration in North West London. The SNOMED codes used are available in Additional File 2 (note that due to updates in the highest-risk criteria between this study and the previous descriptive analysis,^19^ the SNOMED codes used were also updated).

As per the inclusion criteria, patients were required to be non-hospitalised at the time of sotrovimab treatment; to be considered non-hospitalised at the time of treatment, patients must not have had an inpatient hospital visit (event from admission to discharge) starting on or before the date of treatment, unless the visit was a day case (in the NHS, a day case is a planned elective admission without a planned overnight stay, used to administer treatments under medical supervision or to conduct minor procedures) or the visit did not incur an overnight stay.

Patients were excluded if they received more than one COVID-19 treatment (sotrovimab, nirmatrelvir/ritonavir, molnupiravir or remdesivir) in an outpatient setting before the index date, or were diagnosed with COVID-19 while hospitalised.

#### Data analysis

Patient characteristics were recorded, including age, sex, ethnicity, vaccination status and comorbidity history. Cohorts were described in relation to ‘highest-risk’ conditions which made patients eligible for early treatment with sotrovimab and antiviral therapies, as mentioned above, and other high-risk conditions which may make the patient susceptible to adverse outcomes from COVID-19 (see Table 1 for highest- and high-risk comorbidities).

**Table 1.** High- and highest-risk conditions criteria.

| Highest-risk conditions | High-risk conditions |
| --- | --- |
| Down's syndrome | Age ≥70 years |
| Solid cancer | Long-term respiratory conditions |
| Haematological disease and stem-cell transplant recipients | Chronic heart disease |
| Advanced renal disease | Chronic kidney disease |
| Liver disease | Chronic liver disease |
| IMID | Chronic neurological condition |
| Immune deficiencies | Diabetes |
| HIV/AIDS | Weakened immune system caused by medical condition or medication |
| Solid organ transplant | Obesity (class III) |
| Rare neurological conditions | Pregnancy |
|  | Severe respiratory conditions |
|  | Rare disease and inborn errors of metabolism |

AIDS, acquired immune deficiency syndrome; HIV, human immunodeficiency virus; IMID, immune-mediated inflammatory disease.

Continuous variables (e.g. age) were summarised using mean, standard deviation, median, interquartile range and range. Categorical variables (e.g. sex) were described using frequencies and percentages. Values from ≥1 to <5 were suppressed to protect patient confidentiality and are reported as n<5, as per our study’s Information Governance and Data Privacy Impact Assessment approvals.

Inverse probability of treatment weighting (IPTW) was used to balance baseline patient characteristics in the treated and untreated cohorts. Weights were derived based on propensity scores, which were further used in weighted Cox regression to adjust for measured confounders between the treated and untreated cohorts. Propensity scores (probability of treatment based on baseline covariates) were obtained using logistic regression or gradient boosting machine models. Propensity score models were used to predict the probability of treatment based on the following covariates: age, gender, time period of COVID-19 diagnosis (i.e. Omicron BA.1, BA.2 or BA.5, as defined above), presence of renal disease (binary), presence of multiple highest-risk conditions (≥2, binary), presence of high-risk conditions (binary), solid-organ transplant (binary), COVID vaccination status (binary), time since vaccination and ethnicity (a full list of variables and models is included in Additional File 1, Table S1). To obtain an appropriate estimation of the variance of the treatment effect and better control the type I error rate, inverse probability of treatment weights were stabilised.^23^ The balance in baseline characteristics between weighted treated and untreated groups was assessed using standardised differences.

Cox proportional hazards models with stabilised weights were performed to assess the hazard ratio (HR) of COVID-19-related hospitalisation and/or COVID-19-related death among the overall cohort and the patient subgroups (Omicron subvariant prevalence periods, age <65 years and ≥65 years, and patients with renal dysfunction). Covariates not balanced after weighting (standardised differences >0.1) were included in the Cox proportional hazards model. IPTWs and accordingly doubly robust estimation was performed separately for each Cox model. Analyses were conducted using R version 4.2.1 and the following packages: twang 2.5, cobalt 4.4.1, xtable 1.8-4, survey 4.1-1, stringr 1.4.1, WeightIt 0.13.1, stats 4.2.1, survminer 0.4.9, survival 3.3-1, powerSurvEpi 0.1.3, Data pre-processing was performed using Python 3.9.5 with packages Pandas 1.3.4 and Numpy 1.21.

Patients without evidence of at least one of the NHS highest-risk criteria for receiving early treatment were excluded from the main analysis. However, in our previous descriptive analysis, we observed that a high proportion of those prescribed sotrovimab (39.2%) did not have a code for a highest-risk condition.^19^ Therefore, we performed an exploratory analysis whereby a SNOMED code (1300561000000107; high-risk category for developing complication from coronavirus disease 19 caused by severe acute respiratory syndrome coronavirus 2 infection) was used to identify patients who were identified as appropriate for ‘shielding’ during the early phase of the pandemic.

## Results

### Patient demographics and baseline characteristics

The analysis included 5,790 patients, 599 (10.3%) of whom were treated with sotrovimab and 5,191 (89.7%) who were eligible highest-risk untreated patients (Table 2). A total of 2,946 patients were diagnosed during period 1 (173 sotrovimab-treated, 2,773 untreated), 1,978 were diagnosed during period 2 (285 sotrovimab-treated, 1,693 untreated) and 866 were diagnosed during period 3 (141 sotrovimab-treated, 725 untreated).

**Table 2.** Patient characteristics.

|  | **Sotrovimab**<br>**(n=599)** | **Untreated**<br>**(n=5,191)** |
| --- | --- | --- |
| **Age** |  |  |
| Mean (SD) | 57.4 (15.6) | 52.5 (17.6) |
| Median (Q1–Q3) | 58 (46–70) | 53 (40–65) |
| Age group ≥65 years, n (%) | 211 (35.2) | 1,302 (25.1) |
| **Female sex, n (%)** | 303 (50.6) | 2,459 (47.4) |
| **Ethnicity, n (%)** |  |  |
| White | 309 (54.3) | 2,556 (49.2) |
| Asian/Asian British | 136 (23.9) | 1,167 (22.5) |
| Black/Black British | 69 (12.1) | 768 (14.8) |
| Mixed | 18 (3.2) | 189 (3.6) |
| Other | 37 (6.5) | 277 (5.3) |
| Unknown | 30 (5.0) | 234 (4.5) |
| **Vaccination status, n (%)** |  |  |
| Fully<sup>a</sup> | 560 (93.5) | 4,536 (87.4) |
| Not fully | 39 (6.5) | 655 (12.6) |
| **Time since last vaccination (days)** |  |  |
| Mean (SD) | 131.6 (90.3) | 127.2 (89.3) |
| Median (Q1–Q3) | 112 (66–165) | 97 (49–167) |
| **Time to treatment (days)** |  |  |
| Mean (SD) | 1.9 (1.6) | 1.8 (1.6) |
| Median (Q1–Q3) | 1 (1–2) | 1 (1–2) |
| **Highest-risk conditions, n (%)** |  |  |
| Down's syndrome | 6 (1.0) | 138 (2.7) |
| Solid cancer | 72 (12.0) | 487 (9.4) |
| Haematological disease and stem-cell transplant recipients | 71 (11.9) | 400 (7.7) |
| Renal disease | 254 (42.4) | 1,094 (21.1) |
| Liver disease | 41 (6.8) | 487 (9.4) |
| IMiD | 45 (7.5) | 512 (9.9) |
| Immune deficiencies | 75 (12.5) | 1,250 (24.1) |
| HIV/AIDS | 60 (10.0) | 1,149 (22.1) |
| Solid-organ transplant | 72 (12.0) | 244 (4.7) |
| Rare neurological conditions | 81 (13.5) | 797 (15.4) |
| **Number of highest-risk conditions, n (%)** |  |  |
| 1 | 446 (74.5) | 3,887 (74.9) |
| 2 | 129 (21.5) | 1,252 (24.1) |
| 3+ | 24 (4.0) | 52 (1.0) |
| **High-risk conditions, n (%)** |  |  |
| Age ≥70 years | 150 (25.0) | 943 (18.2) |
| Long-term respiratory conditions | 134 (22.4) | 1,116 (21.5) |
| Chronic heart disease | 336 (56.1) | 1,810 (34.9) |
| Chronic kidney disease | 155 (25.9) | 769 (14.8) |
| Chronic liver disease | 60 (10.0) | 572 (11.0) |
| Diabetes | 169 (28.2) | 1,024 (19.7) |
| Weakened immune system caused by medical condition or medication | 122 (20.4) | 780 (15.0) |
| Obesity (class III) | 14 (2.3) | 94 (1.8) |
| Pregnancy | 8 (1.3) | 265 (5.1) |
| Rare disease and inborn errors of metabolism | 8 (1.3) | 265 (5.1) |
| **Number of high-risk conditions, n (%)** |  |  |
| 0 | 103 (17.2) | 1,617 (31.2) |
| 1 | 123 (20.5) | 1,177 (22.7) |
| 2–3 | 263 (43.9) | 1,705 (32.8) |
| 4+ | 110 (18.4) | 692 (13.3) |

<sup>a</sup>Minimum complete vaccination schedule plus at least one booster.
AIDS, acquired immune deficiency syndrome; COVID-19, coronavirus disease 2019; HIV, human immunodeficiency virus; IMID, immune-mediated inflammatory disease; IQR, interquartile range; SD, standard deviation.

Patients aged ≥65 years accounted for 35.2% (n=211/599) of the sotrovimab-treated group and 25.1% (n=1,302/5,191) of untreated patients. A high percentage of patients treated with sotrovimab had renal disease (42.4%, n=254/599 vs 21.1%, n=1,094/5,191 of untreated patients), while lower percentages were reported for other highest-risk comorbidities (Table 2). A high percentage of sotrovimab-treated patients had high-risk comorbidities such as chronic heart disease (56.1%, n=336/599), chronic kidney disease (25.9%, n=155/599) and diabetes (28.2%, n=169/599). Among untreated patients, 34.9% (n=1,810/5,191) had chronic heart disease, 14.8% (n=769/5,191) had chronic kidney disease and 19.7% (n=1,024/5,191) had diabetes. The proportion of patients with at least four high-risk comorbidities was 18.4% (n=110/599) among those treated with sotrovimab and 13.3% for untreated patients (n=692/5,191). The proportion of patients categorised as fully vaccinated (minimum complete vaccination schedule plus at least one booster) was 93.5% (n=560/599) in the sotrovimab group and 87.4% (n=4,356/5,191) in the untreated group.

### Clinical outcomes

After weighting, the time period of COVID-19 diagnosis covariate remained unbalanced between the treated and untreated cohorts and was therefore included in all weighted Cox proportional hazards models (Additional File 1, Table S2).

In the sotrovimab-treated cohort, all-cause and COVID-19-related hospitalisations were experienced by 7.2% (n=43/599) and 1.2% (n=7/599) of patients, respectively. Fewer than five patients died within one month of index (Table 3). In the untreated cohort, all-cause and COVID-19-related hospitalisations were experienced by 5.2% (n=270/5,191) and 1.7% (n=90/5,191) of patients, respectively. Within one month of index, 22 patients (0.4%) died (Table 3).

**Table 3.** HRs for IPTW weighted^a^ Cox proportion hazard for study outcomes.

| Clinical outcomes | Sotrovimab | Untreated |
| --- | --- | --- |
| Overall cohort, n | 599 | 5,191 |
| Compound (hospitalisation or death) |  |  |
| HR | 0.50 |  |
| 95% CI | 0.24, 1.06 |  |
| p-value | 0.07 |  |
| COVID-19-related hospitalisation |  |  |
| Patients with event, n (%) | 7 (1.2) | 90 (1.7) |
| HR | 0.43 |  |
| 95% CI | 0.18, 1.00 |  |
| p-value | 0.051 |  |
| Death |  |  |
| Patients with event, n (%) | <5 | 22 (0.4) |
| HR | 0.71 |  |
| 95% CI | 0.16, 3.20 |  |
| p-value | 0.65 |  |
| Patients aged <65 years, n | 388 | 3,889 |
| Compound (hospitalisation or death) |  |  |
| HR | 0.79 |  |
| 95% CI | 0.34, 1.85 |  |
| p-value | 0.58 |  |
| COVID-19-related hospitalisation |  |  |
| Patients with event, n (%) | 6 (1.5) | 47 (1.2) |
| HR | 0.70 |  |
| 95% CI | 0.28, 1.75 |  |
| p-value | 0.44 |  |
| Death |  |  |
| Patients with event, n (%) | <5 | <5 |
| HR | 1.98 |  |
| 95% CI | 0.21, 18.18 |  |
| p-value | 0.55 |  |
| Patients aged ≥65 years, n | 211 | 1,302 |
| Compound (hospitalisation or death) |  |  |
| HR | 0.25 |  |
| 95% CI | 0.06, 1.12 |  |
| p-value | 0.07 |  |
| COVID-19-related hospitalisation |  |  |
| Patients with event, n (%) | <5 | 43 (3.3) |
| HR | 0.11 |  |
| 95% CI | 0.02, 0.82 |  |
| p-value | 0.03 |  |
| Death |  |  |
| Patients with event, n (%) | <5 | 19 (1.5) |
| HR | 0.55 |  |
| 95% CI | 0.07, 4.05 |  |
| p-value | 0.55 |  |
| Patients without renal disease, n | 345 | 4,097 |
| Compound (hospitalisation or death) |  |  |
| HR | 0.55 |  |
| 95% CI | 0.21, 1.47 |  |
| p-value | 0.24 |  |
| COVID-19-related hospitalisation |  |  |
| Patients with event, n (%) | <5 | 53 |
| HR | 0.55 |  |
| 95% CI | 0.19, 1.62 |  |
| p-value | 0.28 |  |
| Death |  |  |
| Patients with event, n (%) | <5 | 9 |
| HR | 0.54 |  |
| 95% CI | 0.07, 4.27 |  |
| p-value | 0.56 |  |
| Patients with renal disease, n | 254 | 1,094 |
| Compound (hospitalisation or death) |  |  |
| HR | 0.28 |  |
| 95% CI | 0.09, 0.89 |  |
| p-value | 0.031 |  |
| COVID-19-related hospitalisation |  |  |
| Patients with event, n (%) | <5 | 37 |
| HR | 0.18 |  |
| 95% CI | 0.05, 0.62 |  |
| p-value | 0.007 |  |
| Death |  |  |
| Patients with event, n (%) | <5 | 13 |
| HR | 0.51 |  |
| 95% CI | 0.07, 3.89 |  |
| p-value | 0.52 |  |

<sup>a</sup>IPTW included patient age, gender, time period of index, presence of renal disease, presence of multiple highest-risk conditions, vaccination status, days since last vaccination and ethnicity. Cox proportion hazard additionally adjusted for multiple highest-risk conditions, age and ethnicity.
CI, confidence interval; HR, hazard ratio; IPTW, inverse probability of treatment weighting.

The IPTW HRs for COVID-19 hospitalisation or death, COVID-19 hospitalisation and death are shown in Figure 1 and Table 3. Compared with no treatment, the risk of COVID-19 hospitalisation was 57% lower (HR=0.43; 95% confidence interval [CI] 0.18, 1.00; p=0.051) with sotrovimab treatment. Similarly, the risk of COVID-19 hospitalisation or death was 50% lower (HR=0.50; 95% CI 0.24, 1.06; p=0.07) with sotrovimab. The event rate for death was too low for conclusions to be drawn.

**Figure 1.**
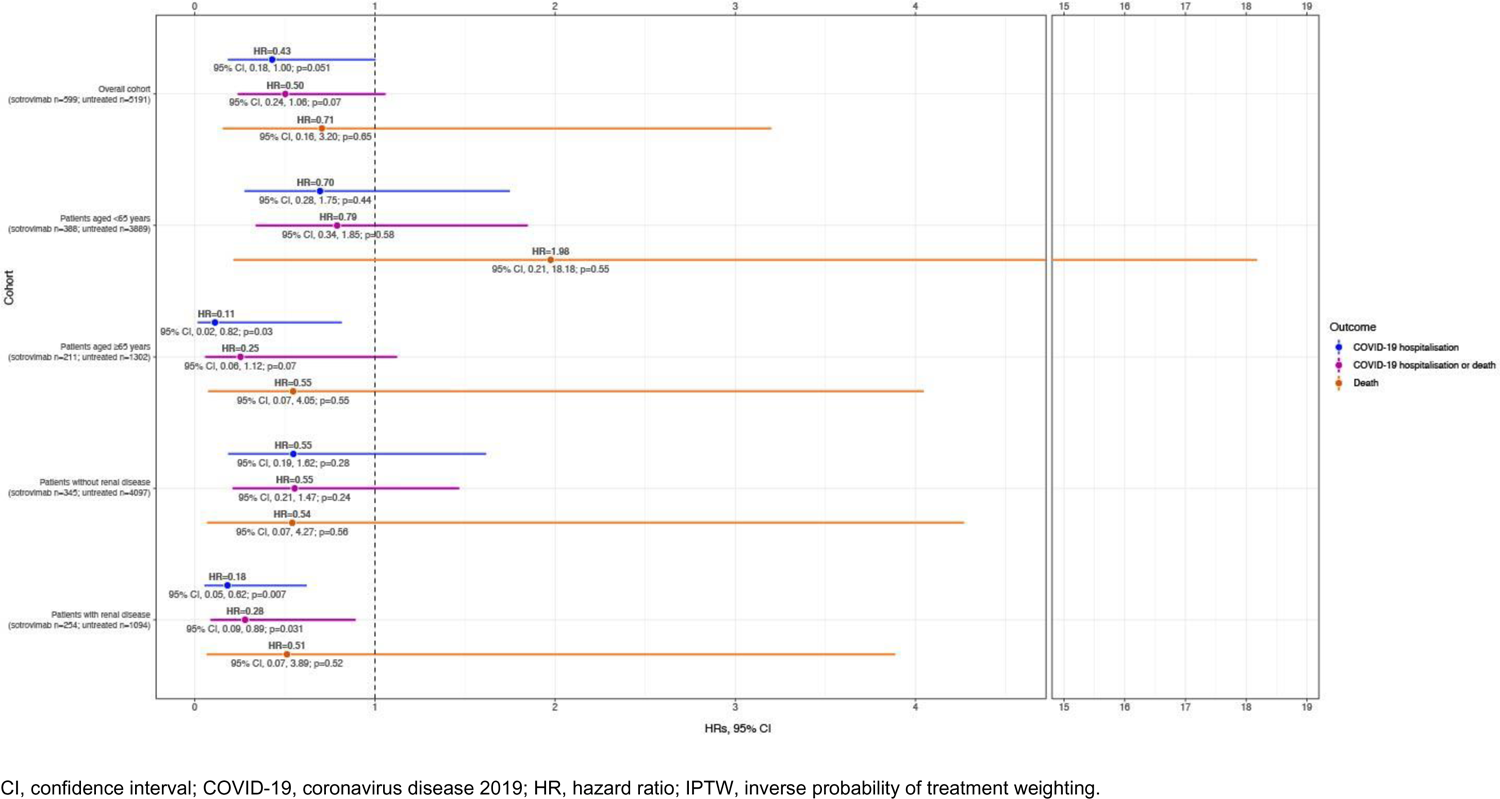
IPTW Cox proportional HRs for COVID-19 hospitalisation and/or death in sotrovimab-treated compared with untreated patients (entire cohort, n=5,790)

### Subgroup analyses

Among patients aged <65 years, COVID-19-related hospitalisations were experienced by 1.5% (n=6/388) of those treated with sotrovimab and 1.2% (n=47/3,889) of those untreated. Fewer than five patients in each cohort died within one month of the index date (Table 3).

The IPTW HR of COVID-19 hospitalisation or death was 0.79 (95% CI 0.34, 1.85; p=0.58) for sotrovimab compared with no treatment (Figure 1). The IPTW HRs for COVID-19 hospitalisation and death in patients aged <65 years were 0.70 (95% CI 0.28, 1.75; p=0.44) and 1.98 (95% CI 0.21, 18.18; p=0.55), respectively (Figure 1; Table 3). The event rate for death was too low for conclusions to be drawn.

Among patients aged ≥65 years, COVID-19-related hospitalisations were experienced by fewer than five of the 211 patients treated with sotrovimab and 3.3% (n=43/1,302) of those untreated. Deaths within one month of index were reported for fewer than five sotrovimab-treated patients and 1.5% (n=19/1,302) of untreated patients (Table 3). Sotrovimab treatment was associated with a statistically significant 89% reduction in the risk of COVID-19 hospitalisation compared with no treatment (HR=0.11; 95% CI 0.02, 0.82; p=0.03) (Figure 1). IPTW HRs for composite COVID-19 hospitalisation or death and death as a single endpoint were 0.25 (95% CI 0.06, 1.12; p=0.07) and 0.55 (95% CI 0.07, 4.05; p=0.55), respectively (Figure 1; Table 3).

Among patients without renal disease, none of the IPTW HRs were statistically significant, although all HRs were <1 (Figure 1). However, sotrovimab treatment was associated with a statistically significant 72% reduction in the risk of COVID-19 hospitalisation or death (HR=0.28; 95% CI 0.09, 0.89; p=0.031) among patients with renal disease compared with no treatment (Figure 1, Table 3). The risk of COVID-19 hospitalisation was also significantly lower by 82%, following sotrovimab treatment compared with the untreated group (HR=0.18; 95% CI 0.05, 0.62; p=0.007). As above, the event rate for death was too low for conclusions to be drawn.

In period 1, sotrovimab treatment was associated with a statistically significant 75% reduction in the risk of COVID-19 hospitalisation or death compared with the untreated group (HR=0.25; 95% CI 0.07, 0.89; p=0.032) (Table 4). In periods 2 and 3, the IPTW HRs of COVID-19 hospitalisation or death were 0.53 (95% CI 0.14, 2.00; p=0.35) and 0.78 (95% CI 0.23, 2.69; p=0.69), respectively, for sotrovimab treatment compared with the untreated group (Table 4).

**Table 4.** HRs of treated versus untreated for IPTW weighted^a^ Cox proportion hazard for study outcomes.

|  | **Period 1<sup>b</sup>**<br>**(n=2,946)** | **Period 2<sup>c</sup>**<br>**(n=1,978)** | **Period 3<sup>d</sup>**<br>**(n=866)** |
| --- | --- | --- | --- |
| **Compound (hospitalisation or death)** |  |  |  |
| HR | 0.25 | 0.53 | 0.78 |
| 95% CI | 0.07, 0.89 | 0.14, 2.00 | 0.23, 2.69 |
| p-value | 0.032 | 0.35 | 0.69 |
| **Hospitalisation** |  |  |  |
| HR | - | 0.51 | 0.60 |
| 95% CI | - | 0.09, 2.72 | 0.14, 2.62 |
| p-value | - | 0.43 | 0.49 |
| **Death** |  |  |  |
| HR | - | 0.59 | 1.04 |
| 95% CI | - | 0.07, 4.75 | 0.11, 9.68 |
| p-value | - | 0.62 | 0.97 |

Cox models could not be solved for hospitalisation and death in Period 1 (likely due to multicollinearity).
<sup>a</sup>IPTW included patient age, gender, time period of index, presence of renal disease, presence of multiple highest-risk conditions, vaccination status, days since last vaccination and ethnicity. Cox proportion hazard additionally adjusted for multiple highest-risk conditions, age and ethnicity.
<sup>b</sup>Patients with an index date of 1 December 2021–12 February 2022.
<sup>c</sup>Patients with an index date of 13 February 2022–31 May 2022.
<sup>d</sup>Patients diagnosed between 1 June 2022–31 July 2022.
CI, confidence interval; HR, hazard ratio; IPTW, inverse probability of treatment weighting.

### Exploratory analysis

A total of 21,320 patients diagnosed with COVID-19 were identified as being appropriate for ‘shielding’ (evidence of a shielding SNOMED code, as outlined in the Methods) during the study period. Of these patients, 17,921 (84.1%) had a shielding code but no specific highest risk-related diagnosis and 3,399 (15.9%) had both. Of those patients identified as being appropriate for ‘shielding’, 692 (3.2%) patients received sotrovimab and 20,628 (96.8%) were untreated. After adding the “shielding-code only” patients to those who also had highest risk diagnostic codes, COVID-19-related hospitalisations were experienced by 1.4% (n=10/692) of patients treated with sotrovimab and 0.9% (n=184/20,628) of untreated patients. Death within one month of index was reported for fewer than five sotrovimab-treated patients, and 0.4% of untreated patients (n=82/20,628). The IPTW HR for composite COVID-19 hospitalisation or death was 1.14 (95% CI 0.48, 2.66; p=0.77) for sotrovimab compared with no treatment; for COVID-19 hospitalisation alone the IPTW HR was 1.46 (95% CI 0.59, 3.57; p=0.41). Sotrovimab treatment was associated with an 82% lower risk of death alone compared with the untreated group (HR=0.18; 95% CI 0.04, 0.73; p=0.02).

## Discussion

This retrospective study assessed the effectiveness of sotrovimab compared with no early COVID-19 treatment in non-hospitalised, highest-risk patients with COVID-19 in North West London, using data from the Discover dataset (one of Europe’s largest linked longitudinal datasets). We previously reported descriptive results for a similar cohort of patients, which were used to confirm the feasibility of this comparative effectiveness analysis.^19^

There was evidence that sotrovimab treatment was associated with a significant reduction in the risk of COVID-19 hospitalisation in vulnerable groups compared with no treatment. An 89% decrease in the hazard of COVID-19-related hospitalisation was observed for patients aged ≥65 years who were treated with sotrovimab (p=0.03). Further, decreases in the hazards of both COVID-19 hospitalisation or death and COVID-19 hospitalisation were observed for patients with renal disease (72% risk reduction [p=0.031] and 82% risk reduction [p=0.007], respectively), who are at especially high risk of severe COVID-19 outcomes.^24^ Similarly, a study conducted among patients on renal replacement therapy found that sotrovimab was associated with a 65% lower risk of 28-day COVID-19-related hospitalisation and/or death than molnupiravir (HR=0.35; 95% CI 0.17, 0.71; p=0.004).^25^

For the overall cohort analysis, the risk of hospitalisation following sotrovimab treatment was reduced by 57% (p=0.051), and the composite hazard of COVID-19 hospitalisation or death by 50% (p=0.07), although these values did not reach statistical significance. Validation of these results on a larger scale would be valuable. Our results are similar to those of a recent United States (US) retrospective analysis of patients diagnosed with COVID-19 during the Delta and early Omicron waves, which reported that sotrovimab was associated with 55% lower risk of 30-day all-cause hospitalization or mortality versus no mAb treatment (p<0.001).^26^ Additionally, a further US study reported a 70% risk reduction in 30-day hospitalisation or mortality among patients treated with sotrovimab versus no treatment during BA.1 predominance.^27^ In the COMET-ICE randomised clinical trial, the risk of all-cause >24-hour hospitalisation or death was reduced by 79% for sotrovimab compared with placebo.^11^ However, it should be noted that the current study was a real-world study, and therefore more likely to include an older population with more comorbidities and greater ethnic diversity. Furthermore, the COMET-ICE trial was conducted whilst the original ‘wild-type’ variant was predominant, rather than the Omicron variants of concern predominant during this study.

For sotrovimab-treated patients, we also report a reduced risk of hospitalisation or death during BA.1 predominance and non-significant trends for reduced risk during BA.2 and BA.5, likely due to a low event rate and small sample size. Harman *et al*. previously reported low proportions of hospital admissions between sequencing-confirmed Omicron BA.1 and BA.2 cases treated with sotrovimab.^28^ Furthermore, a retrospective cohort study using data from the Hospital Episode Statistics database in England reported low levels of COVID-19-attributable hospitalisations and deaths in patients presumed to be treated with sotrovimab (based on NHS data showing that 99.98% of COVID-19-mAb-treated individuals received sotrovimab during the study period), with no significant differences in hospitalisation rates during Omicron BA.1, BA.2 or BA.5 predominance.^29^ Zheng *et al*. used the OpenSAFELY platform to evaluate the comparative effectiveness of sotrovimab and molnupiravir for prevention of severe COVID-19 outcomes between 16 December 2021 and 10 February 2022. They reported that 0.96% of sotrovimab-treated patients had a COVID-19-attributable hospitalisation or death within 28 days of treatment, compared with 2.05% of molnupiravir-treated patients. Cox proportional hazards models showed that sotrovimab was associated with a lower risk than molnupiravir (HR=0.54; 95% CI 0.33, 0.88; p=0.01).^30^ A further study by Zheng *et al*. showed a comparable risk of severe COVID-19 outcomes between patients treated with sotrovimab and nirmatrelvir/ritonavir during Omicron BA.2 and BA.5 predominance in the UK (HR=1.14; 95% CI 0.62, 2.08; p=0.673).^31^ In another recent study using the OpenSAFELY platform that simulated target trials, HRs for COVID-19 hospitalisation or death within 28 days were 0.76 (95% CI 0.66, 0.89) during BA.1 predominance and 0.92 (95% CI 0.79, 1.06) during BA.2 predominance for sotrovimab-treated versus untreated patients.^32^

In our previous descriptive study, ∼40% of patients receiving sotrovimab did not have evidence in their healthcare records of a highest-risk condition that would make them eligible to receive the therapy.^19^ This was an unexpected result: as a high-cost specialist drug, sotrovimab is only for use in the NHS for a specifically defined group of patients. We therefore performed an exploratory analysis to investigate if we were missing a criterion for identifying eligible patients that might instead be captured in the shielding code. By including patients eligible for ‘shielding’ without documented evidence in the database of at least one of the NHS highest-risk criteria for receiving early treatment, our exploratory analysis may have included more patients who had a better prognosis than were included in the main study analysis. Whilst including only patients with unequivocal evidence of highest-risk criteria reduced the sample size for the main analysis, the untreated cohort used in the exploratory analysis may be less comparable to the sotrovimab-treated cohort.

There are some limitations to this retrospective study which should be noted. Although significant efforts were made to account for confounding factors in the analysis, the influence of unidentified and unmeasured confounders cannot be excluded. One explanation for the lack of statistically significant benefit of sotrovimab observed across the full cohort could be that the sample size and the composite endpoint event rates were too small as hospitalisation rates were decreasing, particularly in later months.^33^ Further, unmeasured confounders such as COVID-19 severity and symptoms at baseline were not recorded. In the untreated cohort, health-seeking behaviours (or lack thereof) may have confounded results; vaccination rate amongst the untreated cohort was significantly lower, and we lack real detail on time interval between symptom onset and formal diagnosis. For instance, as Omicron exposure and vaccination uptake reduced population susceptibility to Delta, there were conceivable employment and social factors which mitigated against testing and the potential requirement to isolate that may have been more apparent in the untreated group.

Whereas patients with certain chronic diseases, such as renal failure, were managed along specific pathways and may have had higher surveillance and facilitated access to treatment. As is common to real-world database analyses, our reporting of patient characteristics and comorbidities is dependent on accurate recording of such data by healthcare practitioners; missing and inaccurate data cannot therefore be ruled out. Identification of an appropriate control may also have impacted results; we observed a large number of patients treated with sotrovimab who had no highest-risk conditions that were used to identify controls. In addition, the Discover dataset is restricted to North West London, so it was not possible to evaluate sub-national geographical trends. Our results may also not be generalisable to non-North West London populations. Finally, the likely SARS-CoV-2 variant was defined by an ecological proxy rather than sequencing-confirmation.

## Conclusions

This retrospective study compared the effectiveness of sotrovimab with no early treatment in non-hospitalised, highest-risk patients with COVID-19 in North West London from December 2021–July 2022. Sotrovimab treatment was associated with a significant reduction in risk of COVID-19 hospitalisation in patients aged ≥65 years and those with renal disease compared with the untreated cohort. For the overall cohort, the risk of hospitalisation following sotrovimab treatment was also lower compared to the untreated group; however, this did not achieve statistical significance (p=0.051). The risk of hospitalisation and/or death was lower for the sotrovimab-treated cohort across all time periods but did not reach significance for periods 2 and 3. Further research with a larger sample size should be considered.

## Supporting information

Additional File 2 - SNOMED codes

## Data Availability

The Discover data that support the findings of this study are available from Imperial College Health Partners via approval from the DRAG under certain restrictions.

## Medical writing, editorial and other assistance

Editorial support (in the form of writing assistance, including preparation of the draft manuscript under the direction and guidance of the authors, collating and incorporating authors’ comments for each draft, assembling tables, grammatical editing and referencing) was provided by Kathryn Wardle of Apollo, OPEN Health Communications, in accordance with Good Publication Practice (GPP) 3 guidelines (www.ismpp.org/gpp-2022), and was funded by GSK and Vir Biotechnology, Inc.

## Authorship

All named authors meet the International Committee of Medical Journal Editors (ICMJE) criteria for authorship for this article, take responsibility for the integrity of the work as a whole and have given their approval for this version to be published.

## Author contributions

MD, MJY, VP, BL, DCG, JDW, SY, BFP, HJB, TK and SJB designed the study; ERG performed inverse probability of treatment weighted survival analysis; MJY performed descriptive analysis; MD, ERG, MJY, BL, DCG, JDW, SY, BFP, EJL, WK, TK and SJB interpreted results. All authors took part in drafting, revising or critically reviewing the manuscript; gave final approval of the version to be published; have agreed on the journal to which the article has been submitted; and agree to be accountable for all aspects of the work.

## Compliance with ethics guidelines

This study was approved by the Discover Data Research Access Group (DRAG) which has derogated powers to approve studies under its overarching approval as a Health Research Database. Ethics approval for the use of the Discover Platform for research was obtained from the West Midlands, Solihull HRA research ethics committee (reference 18/WM/0323, Integrated Research Application System project ID 253449). This study complies with all applicable laws regarding subject privacy. Data were aggregated and counts less than five were suppressed in line with information governance suppression rules. No direct subject contact or primary collection of individual human subject data occurred. Study results were in tabular form and aggregate analyses that omit subject identification; therefore, informed consent, ethics committee or institutional review board approval were not required. Any publications and reports do not include subject identifiers.

