## Additional File 1 for "Comparative effectiveness of sotrovimab versus no treatment in non-hospitalised high-risk patients with COVID-19 in North West London: a retrospective cohort study using the Discover dataset"

**Table S1.** Variables and models used for IPTW weighting

| Analysis | PSM used | Variables included in model |
| --- | --- | --- |
| Overall cohort | GBM | Age, gender, time period, renal disease, multiple high-risk conditions, moderate-risk conditions, vaccination status, days since vaccination, ethnicity |
| Patients aged <65 years | Log regression | Age, gender, time period, renal disease, multiple high-risk conditions, moderate-risk conditions, vaccination status, days since vaccination, ethnicity |
| Patients aged ≥65 years | Log regression | Age, gender, time period, renal disease, multiple high-risk conditions, moderate-risk conditions, vaccination status, days since vaccination, ethnicity |
| Patients without renal disease | Log regression | Age, gender, time period, multiple high-risk conditions, moderate-risk conditions, vaccination status, days since vaccination, ethnicity |
| Patients with renal disease | Log regression | Age, gender, time period, multiple high-risk conditions, moderate-risk conditions, vaccination status, days since vaccination, ethnicity |
| Shielded population | GBM | Age, gender, time period, multiple high-risk conditions, moderate-risk conditions, vaccination status, days since vaccination, ethnicity |
| Period 1 | Log regression | Age, gender, renal disease, multiple high-risk conditions, moderate-risk conditions, vaccination status, days since vaccination, ethnicity |
| Period 2 | Log regression | Age, gender, renal disease, multiple high-risk conditions, moderate-risk conditions, vaccination status, days since vaccination, ethnicity |
| Period 3 | Log regression | Age, gender, renal disease, multiple high-risk conditions, moderate-risk conditions, vaccination status, days since vaccination, ethnicity |

GBM, gradient boosting machine; IPTW, inverse probability of treatment weighting; PSM, propensity score matching.

**Table S2.** IPTW metrics

| Variable | Overall cohort | Patients aged <65 years | Patients aged ≥65 years | Patients without renal disease | Patients with renal disease | Shielded population | Period 1 | Period 2 | Period 3 |
|---|---|---|---|---|---|---|---|---|---|
| Age | 0.08 | 0.08 | 0.03 | 0.03 | -0.00 | -0.01 | 0.11 | 0.07 | -0.01 |
| Gender | | | | | | | | | |
| Female | 0.06 | 0.05 | -0.02 | 0.05 | 0.04 | -0.01 | 0.05 | 0.09 | -0.06 |
| Male | -0.06 | -0.05 | 0.02 | -0.05 | -0.04 | 0.01 | -0.05 | -0.09 | 0.06 |
| Ethnicity | | | | | | | | | |
| Asian/Asian British | 0.01 | 0.07 | 0.03 | 0.07 | 0.04 | -0.05 | 0.04 | -0.02 | 0.03 |
| Black/Black British | -0.08 | -0.03 | 0.03 | -0.07 | 0.03 | -0.07 | -0.16 | -0.02 | 0.05 |
| Mixed | -0.06 | 0.03 | -0.01 | 0.09 | -0.00 | -0.05 | 0.02 | -0.01 | 0.04 |
| White | 0.09 | -0.02 | -0.08 | -0.00 | -0.08 | 0.11 | 0.12 | -0.02 | -0.06 |
| Other | -0.03 | -0.05 | 0.06 | -0.08 | 0.03 | 0.01 | -0.10 | 0.08 | -0.01 |
| Null | 0.00 | -0.01 | 0.05 | 0.00 | -0.00 | -0.03 | 0.03 | 0.03 | -0.01 |
| Time period of COVID-19 diagnosis | | | | | | | | | |
| Period 1 | -0.12 | -0.07 | -0.07 | -0.01 | 0.05 | -0.12 | - | - | - |
| Period 2 | 0.10 | 0.02 | 0.04 | -0.01 | -0.02 | 0.11 | - | - | - |
| Period 3 | 0.03 | 0.08 | 0.03 | 0.03 | -0.04 | 0.03 | - | - | - |
| Renal disease | | | | | | | | | |
| Yes | 0.07 | 0.07 | 0.07 | - | - | 0.05 | 0.03 | -0.02 | -0.04 |
| No | -0.07 | -0.07 | -0.07 | - | - | -0.05 | -0.03 | 0.02 | 0.04 |
| Multiple high-risk conditions | | | | | | | | | |
| Yes | -0.06 | -0.11 | 0.01 | -0.11 | -0.00 | 0.05 | -0.16 | -0.08 | -0.03 |
| No | 0.06 | 0.11 | -0.01 | 0.11 | 0.00 | -0.05 | 0.16 | 0.08 | 0.03 |
| Moderate-risk conditions | | | | | | | | | |
| Yes | 0.06 | 0.02 | 0.01 | 0.01 | 0.03 | -0.02 | -0.06 | 0.02 | -0.04 |
| No | -0.06 | -0.02 | -0.01 | -0.01 | -0.03 | 0.02 | 0.06 | -0.02 | 0.04 |
| Solid-organ transplant recipient | | | | | | | | | |
| Yes | 0.01 | -0.02 | 0.00 | -0.01 | -0.00 | 0.05 | -0.04 | -0.01 | -0.00 |
| No | -0.01 | 0.02 | -0.00 | 0.01 | 0.00 | -0.05 | 0.04 | 0.01 | 0.00 |

| | | | | | | | | | |
|---|---|---|---|---|---|---|---|---|---|
| Vaccination status | | | | | | | | | |
| Yes | 0.03 | -0.09 | -0.06 | -0.10 | -0.07 | 0.00 | -0.04 | -0.03 | -0.07 |
| No | -0.03 | 0.09 | 0.06 | 0.10 | 0.07 | -0.00 | 0.04 | 0.03 | 0.07 |
| Days since last vaccination | -0.06 | 0.09 | 0.03 | 0.07 | 0.08 | -0.09 | -0.02 | 0.02 | 0.10 |

COVID-19, coronavirus disease 2019; IPTW, inverse probability of treatment weighting.

**Figure S1.** Omicron subvariant prevalence in England[34]

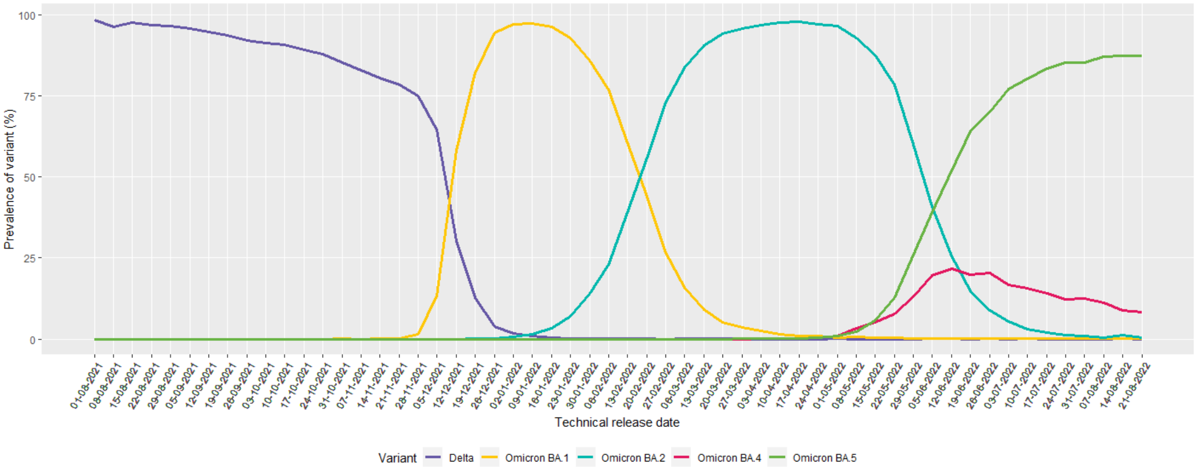

**Figure S2.** Observed time to treatment, and imputed time to treatment in untreated cohort

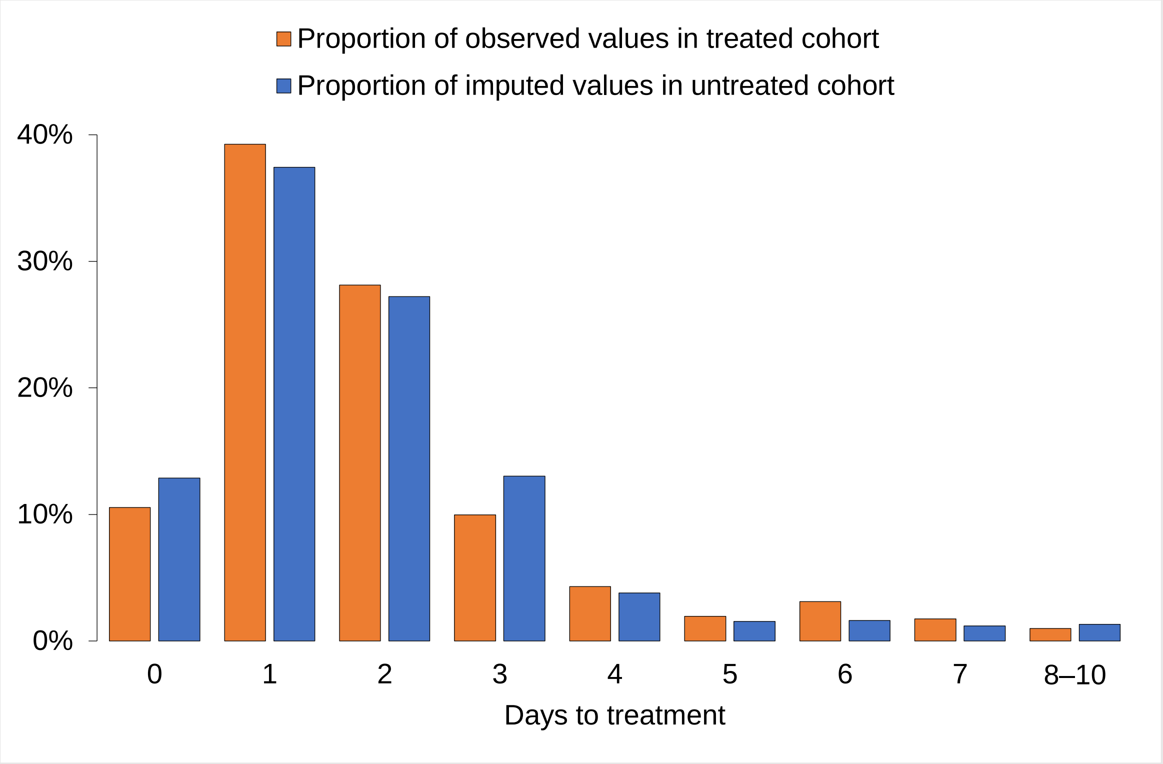
